# Viral Molecular Mimicry Influences the Antitumor Immune Response in Murine and Human Melanoma

**DOI:** 10.1101/2020.09.09.20191171

**Authors:** Jacopo Chiaro, Henna H. E. Kasanen, Thomas Whalley, Cristian Capasso, Mikaela Grönholm, Sara Feola, Karita D. Peltonen, Firas S. Hamdan, Micaela M. Hernberg, Siru Mäkelä, Hanna Karhapää, Paul E. Brown, Beatriz Martins, Manlio Fusciello, Erkko Ylösmäki, Anna S. Kreutzman, Satu M. Mustjoki, Barbara Szomolay, Vincenzo Cerullo

**Affiliations:** Laboratory of Immunovirotherapy, Drug Research Program, Faculty of Pharmacy, University of Helsinki, Viikinkaari 5E, 00790 Helsinki, Finland.; TRIMM, Translational Immunology Research Program, University of Helsinki, Finland.; Hematology Research Unit Helsinki, Department of Hematology, University of Helsinki and Helsinki University Hospital Comprehensive Cancer Center, Haartmaninkatu 8, 00290, Helsinki, Finland.; Department of Clinical Chemistry, University of Helsinki, Helsinki, Finland.; Systems Immunity Research Institute, Cardiff University School of Medicine, Cardiff, United Kingdom.; Department of Oncology, Comprehensive Cancer Center, Helsinki University Hospital and Helsinki University.; Warwick Systems Biology Centre, University of Warwick.; iCAN Digital Precision Cancer Medicine Flagship, University of Helsinki.; HiLIFE Helsinki Institute of Life Science.; Department of Molecular Medicine and Medical Biotechnology and CEINGE, Naples University Federico II, 80131, Naples, Italy.

## Abstract

Molecular mimicry is known to be one of the leading mechanisms by which infectious agents may induce autoimmunity. However, whether a similar mechanism triggers anti-tumor immune response is unexplored, and the role of anti-viral T-cells infiltrating the tumor has remained anecdotal. To address this question, we first developed a bioinformatic tool to identify tumor peptides with high similarity to viral epitopes. Using peptides identified by this tool, we showed that, in mice, viral pre-existing immunity enhanced the efficacy of cancer immunotherapy via molecular mimicry. Specifically, when treated with a cancer vaccine consisting of peptides with a high degree of homology with specific viral peptides, the mice with induced pre-existing immunity to these viral peptides showed significantly better anti-tumor response.

To understand whether this mechanism could partly explain immunotherapy-response in humans, we analyzed a cohort of melanoma patients undergoing PD1 treatment with high IgG titer for Cytomegalovirus (CMV). In this cohort of patients, we showed that high level of CMV-antibodies was associated with a prolonged progression free survival, and found that in some cases PBMCs could cross-react with both melanoma and CMV homologous peptides. Finally, T cell TCR sequencing revealed expansion of the same CD8+ T-cell clones, when PBMCs were pulsed with tumor- or homologous viral peptides.

In conclusion, we have demonstrated that pre-existing immunity and molecular mimicry could explain part of the response observed in immunotherapy. Most importantly, we have developed a tool able to identify tumor antigens and neoantigens based on their similarity to pathogen antigens, in order to exploit molecular mimicry and cross-reactive T-cells in cancer vaccine development.

**One Sentence Summary:** Molecular mimicry can play a role in anti-tumor immune responses and should thus be further exploited in the development of novel cancer treatments.

## Introduction

CD8^+^ T-cells have a key role in the detection and elimination of cells that present abnormal peptides on their surface as a result of viral infection or malignant transformation. Due to the promiscuity of the T-cell receptor (TCR), T-cells recognize a large variety of different targets allowing a relatively small number of T cells to recognize multiple pMHC (peptide:Major Histocompatibility Complex) molecules that can represent a threat^1, 2^. However, a downside of this mechanism is that the immune response directed against a pathogen might result in a potential recognition of self-antigens. This, consequently, can cause a deleterious off-target effect mediated by cross-reactive T-cells, a process known as molecular mimicry^3^. The result of such homology between peptides is a well-established process in the field of autoimmunity, however, it has thus far not been explored in cancer.

It has been thought that the best prognostic marker for successful outcome of immunotherapeutic treatment is the high mutational burden of the tumor and abundant T-cell infiltration, according to the rationale that a tumor that has a high number of mutations will have a higher chance of being recognized and eliminated by infiltrating T-cells^4, 5, 6, 7, 8^. Nevertheless, pivotal studies have shown that qualitative properties of tumor neoantigens might be more important than their quantity. It has been proposed that tumor antigens are more likely to be immunogenic if they resemble infectious-disease-associated antigens, because they are more likely to be recognized by a T-cell^4, 8^. Furthermore, it has been observed that anti-viral T-cells populate the tumor microenvironment^9^, however whether their role is active or not is still unclear.

We have hypothesized that tumors might present peptides that share a high degree of homology with viral peptides and thus might enable cross-reactive T-cells to recognize and promiscuity of the T-cell receptor (TCR), T-cells recognize a large variety of different targets kill tumor cells via molecular mimicry. To assess whether the molecular mimicry is involved in anti-tumor T cell response, we developed a bioinformatic tool called HEX (Homology Evaluation of Xenopeptides) to identify tumor-specific peptides highly similar to viral-derived peptides.

Using several sets of murine and human tumor-specific peptides highly homologous to viral peptides identified by HEX, we herein examine the role of viral pre-existing immunity in cancer anti-tumor T cell response through molecular mimicry.

## Results

### Development of Homologous Evaluation Xenopeptides (HEX) for identification of viral- and tumor-derived peptides with high molecular mimicry

Molecular mimicry is known to be one of the leading mechanisms by which infectious agents may induce autoimmunity^3^, however, whether a similar mechanism drives anti-viral T cells to the tumor and triggers anti-tumor immune response is unexplored and anecdotal. In fact, a tool for a systematic analysis of molecular mimicry between tumor and viral antigens is lacking.

To address this problem and study whether molecular mimicry between viruses and tumors could impact the anti-tumor immune response, we developed HEX, a software that compares the input sequences to a custom database of pathogen-derived protein sequences and selects highly homologous candidate pairs of peptides based on the three following criteria: 1) B-score, that corresponds to the likelihood of the peptides being recognized by a given TCR; 2) the positional weighted alignment score in order to prioritize the similarity in the area of interaction with the TCR; 3) the prediction of the MHC class I binding affinity (Figure 1).

**Figure 1.**
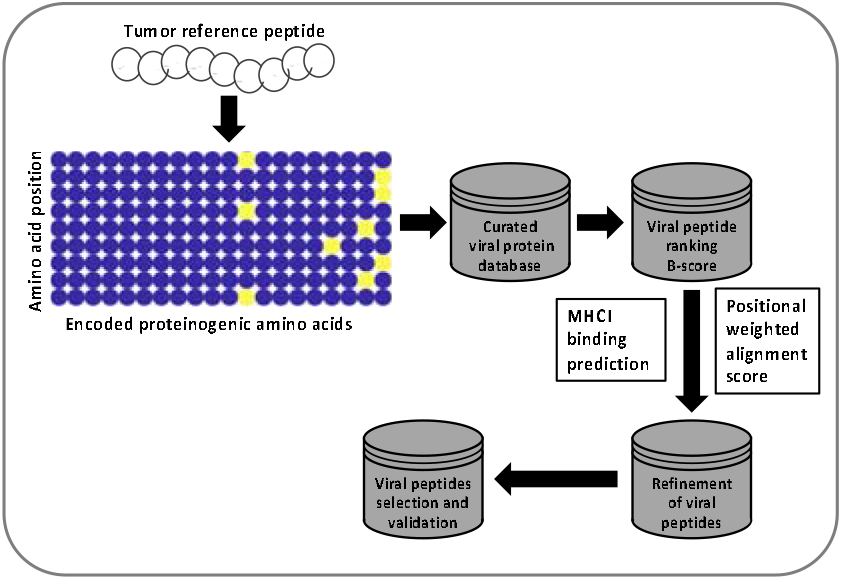
Flowchart of the HEX algorithm. A matrix is generated based on the amino acid composition of a tumor peptide (reference peptide). This matrix is then used to scan the viral database and resulting viral peptides are ranked in order of log-likelihood of recognition (B-score). To each viral peptide is assigned an alignment score and a score for MHC-I binding prediction. The candidate viral peptides are ranked based on the following criteria: MHC-I binding prediction score > alignment score > B-score, and the highest scoring peptides are analyzed experimentally.

To validate the efficiency of the software in selecting homologous peptides with real biological mimicry, we designed an experiment where the tumor growth was followed in mice pre-immunized with viral peptides similar to known tumor antigens. For this experiment, we considered three murine melanoma-associated antigens, that have been applied successfully in a number of vaccination studies: TRP2_180–188_ (tyrosinase-related protein 2), GP100_25–33_ (PMEL; premelanosome protein) and TYR1_208–216_ (tyrosinase 1)^10, 11, 12, 13^. The selected TYR1 sequence was not predicted to bind murine C57BL/6 MHCs, and thus, was considered an “irrelevant peptide”. By HEX analysis, we identified viral peptides that shared high degree of homology with the input tumor epitopes. Pools of 4 viral-derived peptides per each original tumor epitope (Supplementary Table S1) were chosen for further evaluation *in vivo*.

C57BL6 mice were pre-vaccinated with selected viral peptide-pools followed by the tumor engraftment (Figure 2A). Interestingly, all immunized mice showed a reduction in tumor growth compared to mice non-pre-immunized and significant differences between the treatment groups were observed. Particularly, the tumor growth was most reduced in mice immunized with viral peptides homologous to TRP2 or gp100 (Figure 2B).

**Figure 2.**
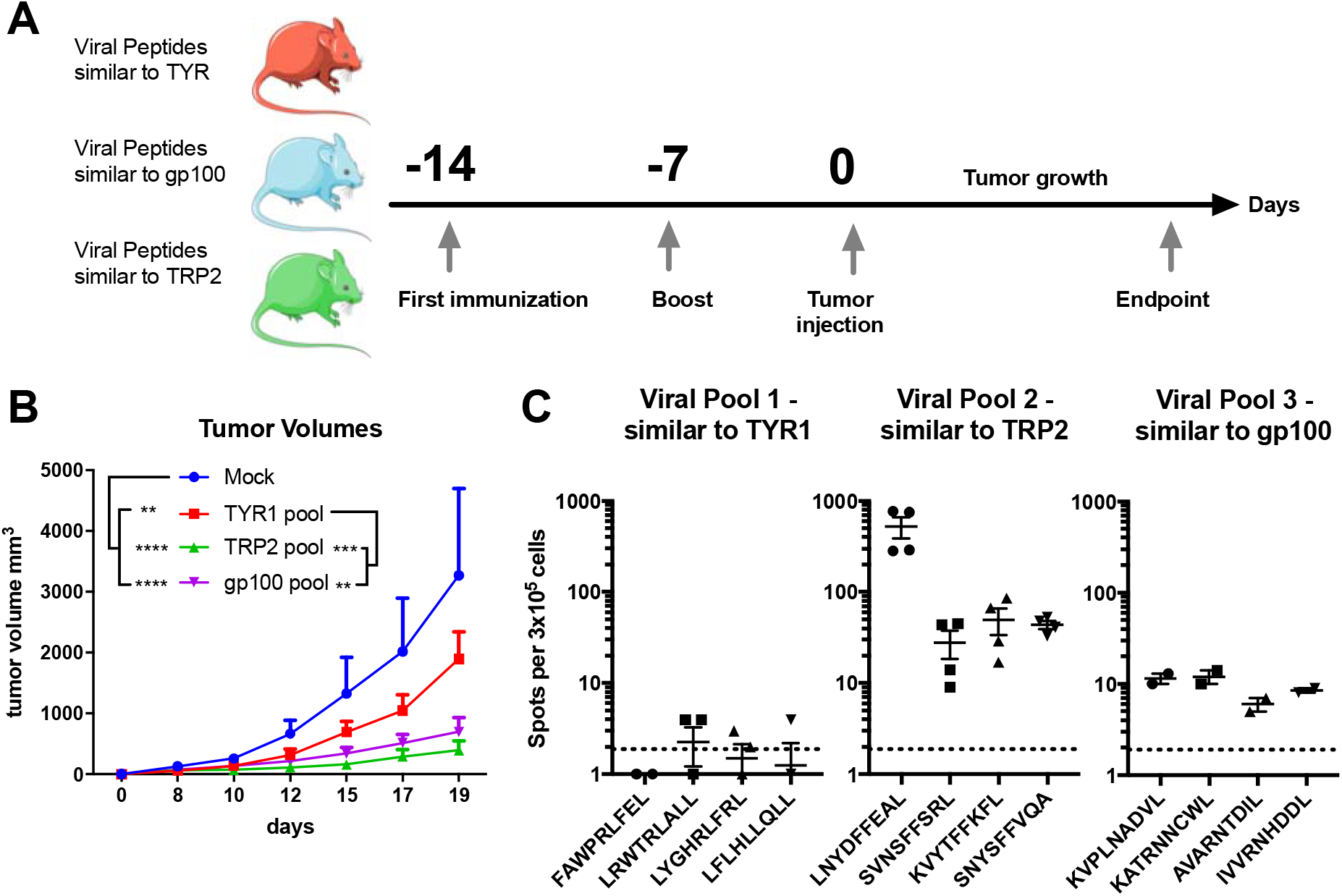
Immunization with viral peptides homologous to tumor antigens slows down tumor growth. Scheme of the animal experiment: To assess if viral peptides similar to tumor peptides can impact tumor growth, 4 groups of C57BL6 mice were formed. A group of naïve mice was used as mock, the other three groups were each immunized with a different pool of viral peptides. The mice were immunized at 2 time points, 14 and 7 days, before the engraftment of the tumor (**A**). After two weeks from the first immunization mice were injected subcutaneously with 3×10^5^murine melanoma B16-OVA cells. After the engraftment, tumor growth was followed by measuring with digital caliper every second day for nineteen days. P value was calculated using Two-Way ANOVA multiple comparison with Tukey’s correction (**B**). Mice were euthanized when the endpoint was reached. The splenocytes of the mice of each group were collected and pooled for an ELISpot assay. Each pool was then pulsed with the respective viral peptides (viral peptides homologous to TYR1, viral peptides homologous to TRP2, viral peptides homologous to GP100) to assess the response to the treatment. The dotted line shows the background produced by the negative control (**C**). The range of p value was labeled with asterisks according to the following criteria: > 0.05 (ns), ≤0.05 (*), ≤0.01 (**), ≤0.001 (***), ≤0.0001 (****).

To further investigate the contribution of the selected viral peptides in the reduction of tumor growth, we collected the splenocytes of the mice at the endpoint for ELISpot assay (Figure 2C). The splenocytes from mice immunized with viral peptide-pools homologous to TRP2 or gp100 fostered higher IFN-γ release compared to the splenocytes from mice immunized with control TYR1-homologous epitopes, in correlation with the outcome of the tumor growth.

We further investigated the reactivity of these splenocytes from the viral pre-immunized mice towards their respective cognate tumor antigen (Figure 3A). When compared side by side only the viral peptides homologous to TRP2 showed higher IFN-γ secretion compared to the cognate tumor peptide (Figure 3B). In addition, we retrospectively assessed the affinities of each tumor antigen and their homologous viral pools to the C57BL/6J MHC class I molecules (Figure 3C) and observed significant correlation with their respective ability to stimulate IFN-γ secretion (Figure 3D). Following the guidelines of the Immune Epitope Database (IEDB) we divided the peptides into High and Low affinity using the threshold of 50nM (http://tools.iedb.org/mhci/help/) and observed that peptides with high MHC-I affinity fostered the production of more INF-γ compared to peptides with low MHC-I affinity (Figure 3E). Taken together, all these results validate the predictive efficiency of HEX to identify homologous peptides with a biologically relevant mimicry.

**Figure 3.**
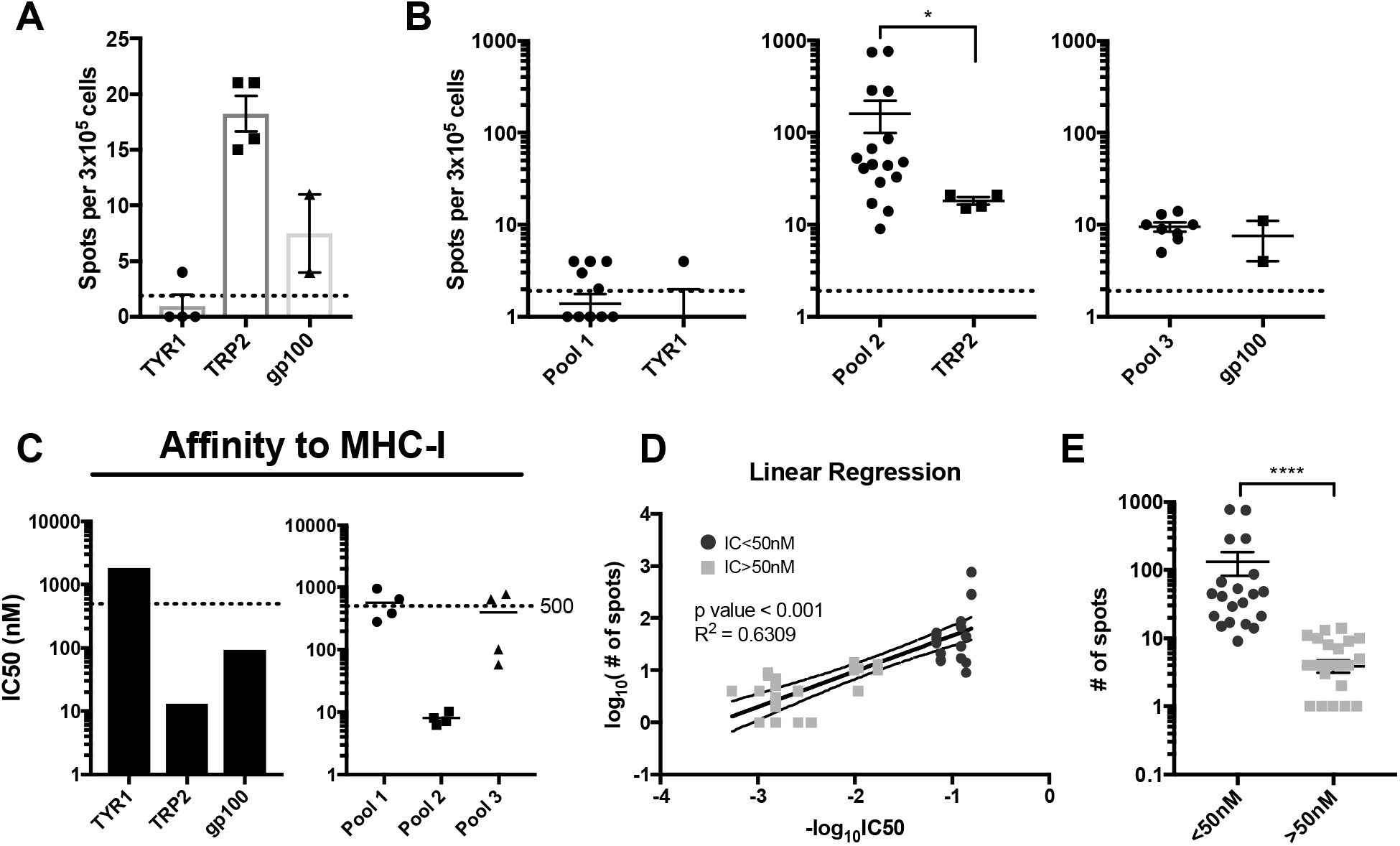
Viral peptides with higher affinity for the MHC are more immunogenic and can elicit stronger cross-reactive response. To assess the response toward the respective original tumor epitope, splenocytes from each group of immunization were pooled together and pulsed with the corresponding tumor peptide (dotted line represent background signal) (**A**). Comparison between the responses elicited by pulsing the splenocytes with the pooled viral peptides and the corresponding original tumor peptide (dotted line represent background signal) (**B**). Predicted affinity of the original tumor and respective similar viral pool of peptides to the murine MHC class I. 50nM was considered as threshold to define peptides with “High-” and “Intermediate/Low Affinity” according to the IEDB guidelines (**C**). Correlation between data from IFN-γ response and predicted affinity (**D**). Stratification of peptides based on their affinity and ability to stimulate IFN-γ production. High affinity peptides (IC50 < 50nM) foster significantly higher production of INF-γ compared to Intermediate/Low affinity peptides (IC50 > 50nM) (**E**). P value was calculated using t test with Mann Whitney correction. The range of p value was represented as follows: > 0.05 (ns), ≤0.05 (*), ≤0.01 (**), ≤0.001 (***), ≤0.0001 (****).

To further validate the efficiency of the HEX selected peptides, we designed another experiment to assess whether the molecular mimicry between viral and tumor antigens could also affect the growth of already established tumors in naïve mice. To this end, mice were engrafted with B16OVA or the more aggressive and immunosuppressive B16F10 tumors and, subsequently, treated with the previously developed vaccine platform PeptiCRAd^14^ consisting of adenoviruses coated with MHC-I restricted peptides (Figure 4A). For this experiment, we utilized the most effective pool of peptides from the previous results. In fact, vaccines were prepared by coating the adenoviruses with the original TRP2_180–188_ epitope (TRP2-PeptiCRAd) or the corresponding pool of viral-derived peptides similar to TRP2 (Viral PeptiCRAd). Intratumoral administration of PeptiCRAd significantly reduced the tumor progression compared to treatment with saline buffer or uncoated-virus both for B16OVA (Figure 4B) and B16F10 tumors (Figure 4D). Mice treated with the original tumor antigen (TRP2) or the viral homologous showed higher therapeutic success rate in both of the tumor models (Figure 4C, 4E). Taken together this first set of experiments suggests that our tool, HEX, is efficient in selecting peptides that share molecular mimicry and this molecular mimicry has a biological relevance.

**Figure 4.**
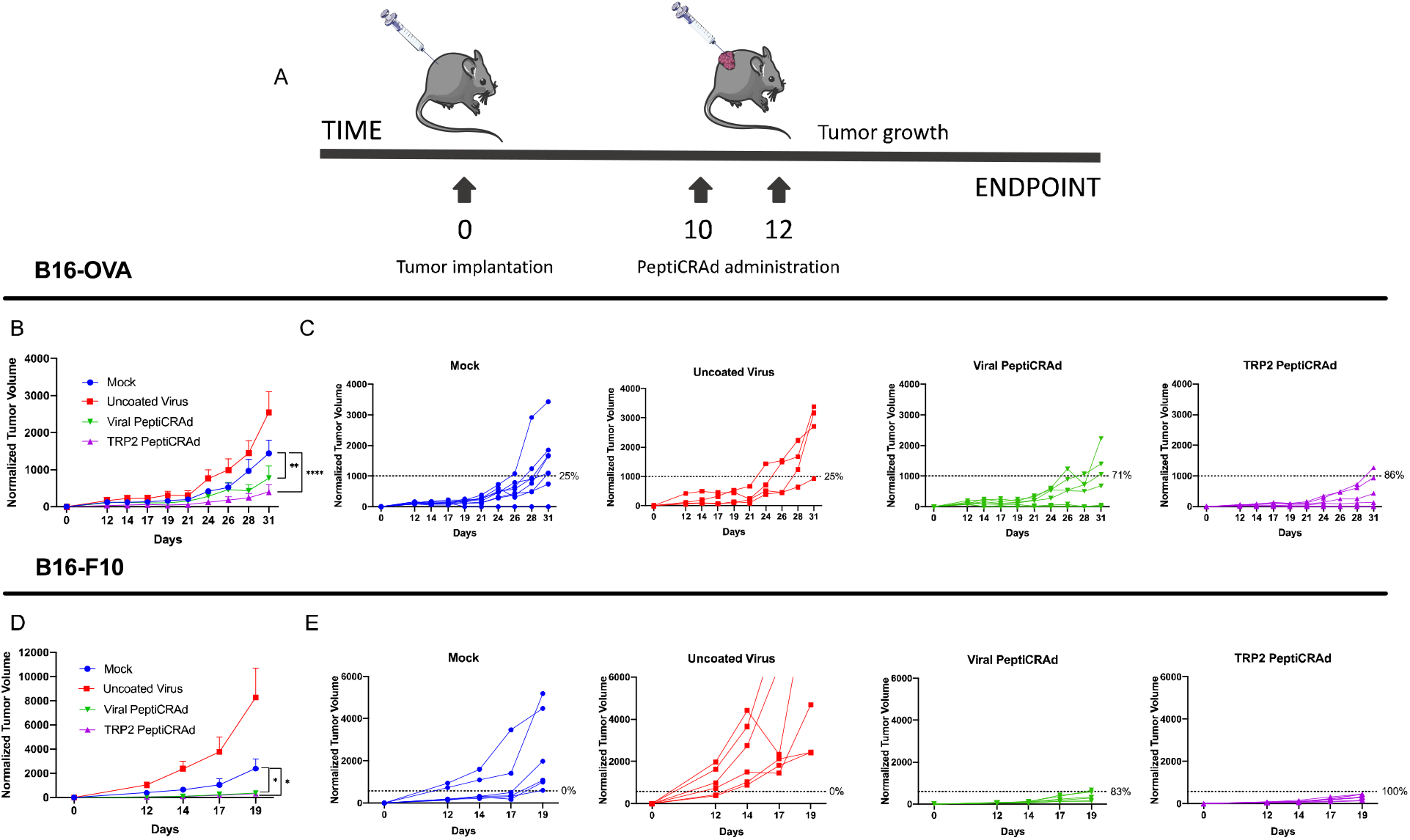
Viral peptides homologous to tumor antigens can reduce tumor growth in already established tumors. Either B16-OVA or B16F10 cells were subcutaneously injected at day 0. As soon the tumors were visible, the mice were treated with either PBS (Mock), Adenovirus alone (Uncoated virus), Adenovirus coated with viral-derived peptides similar to TRP2 (Viral-PeptiCRAd), or Adenovirus coated with TRP2 peptide (TRP2-PeptiCRAd) (**A**). Normalized B16-OVA (**B**) and B16F10 (**D**) tumor volume is shown as mean ± SEM (statistical analysis Two-way ANOVA with multiple comparison). Normalized B16-OVA (**C**) and B16-F10 (**E**) tumor volume curves of individual mice per each group. The dotted line identifies the therapeutic success threshold and represents the median of the normalized volumes measured at the endpoint. *p* value is represented as follow: > 0.05 (ns), ≤0.05 (*), ≤0.01 (**), ≤0.001 (***), ≤0.0001 (****).

### Pre-existing immunity to viruses enhances peptide-based cancer immunotherapy via molecular mimicry

After having established the efficiency of HEX and the biological efficacy of its peptides, we could investigate whether the pre-existing immunity to a given virus would be beneficial when mice are treated with a cancer vaccine based on peptides homologous to that same virus.

To mimic the anti-viral pre-existing immunity, we preimmunized half of a cohort of mice with viral-derived peptides; as before, these viral peptides were selected by HEX to be similar to TRP2, the antigen that gave the best anti-tumor response in previous experiments. After pre-immunization, all the mice were engrafted with the syngeneic B16-OVA and mice that developed palpable tumors were randomized and treated as follows: PBS (Mock), PeptiCRAdgp-100 (no pre-existing immunity for this peptide) and PeptiCRAd-TRP2 (peptide with homologous pre-existing immunity) (Figure 5A). We observed that mice treated with gp100-coated PeptiCRAd displayed no statistically significant difference in tumor volume regardless of their preimmunization status (Supplementary Figure 1A, B). Contrarily, mice preimmunized with peptides similar to TRP2 showed both a better control of tumor growth compared to the naïve group when treated with TRP2-coated PeptiCRAd (Figure 5B) and a higher therapeutic success rate (83% vs 57%) (Figure 5C). At the experimental endpoint, mice were euthanized and flow cytometry analysis was performed on the collected tumors. Interestingly we observed that the anti-viral immunity had conditioned the tumor to become more responsive to the therapy. In fact, we observed an increase of CD8+ and memory CD8+ T cells in the tumor of pre-vaccinated mice, together with their proliferation activity (Ki67+). Interestingly, we also observed a decrease of CD8+CXCR3+ population in pre-immunized mice, suggesting a more memory phenotype in preimmunized mice (Figure 5D). These results support the hypothesis that antiviral-pre-existing immunity can boost the effect of immunotherapies in an antigen specific fashion if there is enough homology between pre-existing antigens and the antigens used for the therapeutic vaccine.

**Figure 5.**
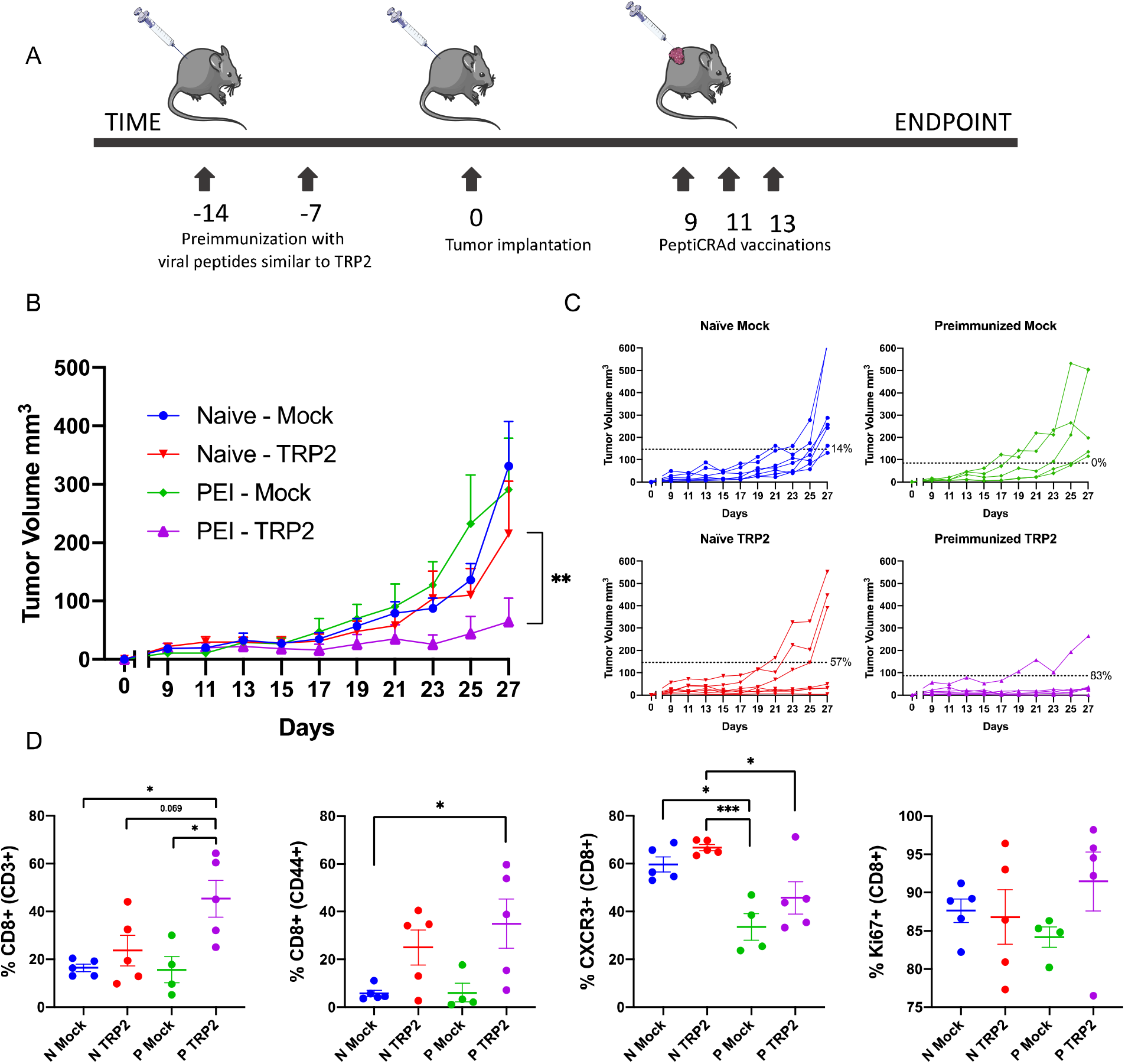
Pre-existing immunity towards viral-derived epitopes mimicking tumors antigens increase efficacy of cancer immunotherapy. Schematic of the treatment pipeline (**A**). Average tumor growth shown as mean ± SEM of each treatment group (**B**). Statistical analysis Two-way ANOVA. Tumor volume curves of individual mice per each treatment group (**C**). The dotted line identifies the threshold of the therapeutic success rate and represents the median of the normalized volumes measured at the endpoint for the Naïve groups and the PEI (pre-immunized) groups respectively. Flow cytometry analysis of the tumors collected at the endpoint (where N stands for Naïve and P stands for pre-immunized) (**D**). p value is represented as follow: > 0.05 (ns), ≤0.05 (*), ≤0.01 (**), ≤0.001 (***), ≤0.0001 (****).

### MHC-I epitope mimicry and cross-reactive T-cells contribute to prolonged survival of anti-PD1 treated metastatic melanoma patients

As demonstrated above, the similarity between tumor antigens and viral-derived antigens can play a crucial role in tumor clearance via cross-reactive T-cells in murine model of cancer. Thus, we wanted to study whether molecular mimicry could also play a significant role in human cancer patients. To this end we studied a cohort of metastatic melanoma patients undergoing treatment with anti-PD1 monotherapy. We collected peripheral blood samples from patients before initiation of therapy, after one and three months and tested their serological status for CMV and Epstein-Barr Virus (EBV) at each timepoint. Using Cox regression, the association of pre-treatment serum CMV and EBV specific IgG levels to progression free survival (PFS) was analyzed. Our results indicated that patients with pretreatment titer of serum anti-CMV IgG higher than the median had significantly longer PFS when compared to patients with anti-CMV IgG levels lower than the median (HR = 0.34, 95% CI: 0.08–1.4, p = 0.04) (Figure 6A). On the contrary, the anti-EBV specific IgG was not associated with prolonged PFS (HR = 0.75 95% CI: 0.2–3.1, p = ns) (Figure 6B).

**Figure 6.**
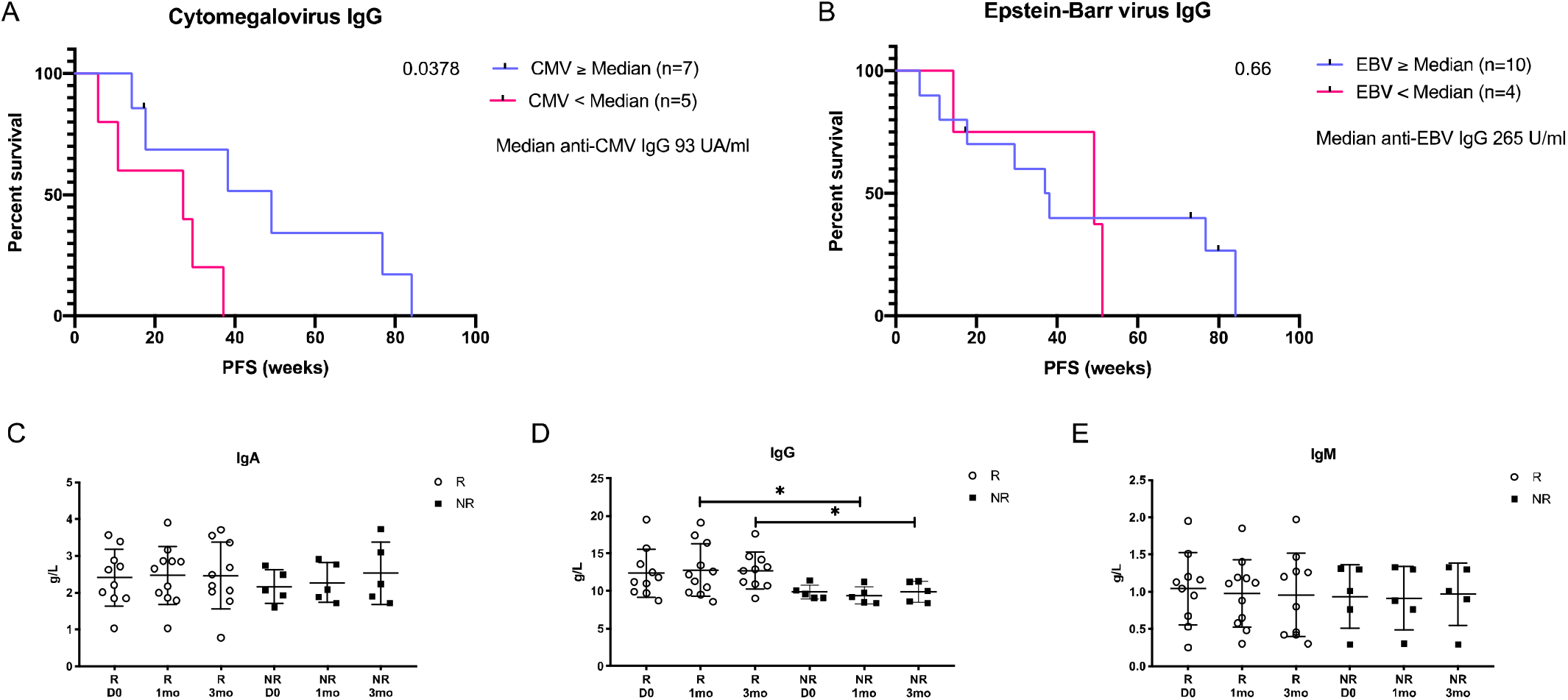
Common viral infections influence prolonged progression-free survival in ICPI-treated metastatic melanoma patients. Kaplan-Meier curve of anti-CMV-IgG high patients (blue line, n = 7) compared to anti-CMV-IgG low (pink line, n = 5) patients (p = 0.05) using Cox regression model. Patients were divided based on their pretreatment anti-CMV-IgG titer as higher, equal (≥) or lower (<) than the median. CMV-seronegative patients were excluded from this analysis. Patients that ended their treatment due to severe adverse effects were censored (black tic marks) (**A**). Kaplan-Meier curve of anti-EBV-IgG high patients (blue line, n = 10) compared to anti-EBV-IgG low (pink line, n = 4) patients (p>0.99) using Cox regression model. Patients were divided based on their pretreatment anti-EBV-IgG titer as higher, equal (≥) or lower (<) the median. EBV-seronegative patients were excluded from this analysis. Patients that ended their treatment due severe adverse effects were censored (black tic marks) (**B**). The immunoglobulin assay of IgA, IgG and IgM serum levels (g/L) between responders (R, white dots) and non-responders (NR, black squares) before initiation (D0) and after one (1mo) and three months (3mo) of ICPI-therapy. The statistical difference between timepoints is calculated with Student’s t-test and the range of p value is labeled with asterisk (*P < 0.05) (**C, D, E**).

To evaluate the more general immunological status of patients, quantitative assessment of IgA, IgG and IgM was performed at each timepoint. The levels of immunoglobulins showed no differences between the responders (R) and non-responders (NR) at pretreatment, indicating that there was no general increased immune-reactive status in patients before initiation of checkpoint inhibitor therapy (Figure 6 C-E). Further, after initiation of anti-PD1 treatment the responders’ IgG levels were significantly higher as compared to non-responders (mean: R_1mo_ 12.8 g/L vs. NR_1mo_: 9.4 g/L, p = 0.02, R_3mo_ 12.7 g/L vs. NR_3mo_: 9.9 g/L) (Figure 5D). Taken together our data indicates that anti-CMV, but not anti-EBV immunity, correlates with prolonged PFS in metastatic melanoma patients undergoing immune-checkpoint inhibitor (ICPI) therapy.

Our findings presented us with a good opportunity to study whether molecular mimicry between CMV and tumor antigens could explain the better prognosis. In fact, we hypothesized that, at least in some patients, cross-reactive T cells between known melanoma antigens and CMV might explain the better PFS. To this end, we selected a pool of melanoma-associated proteins^15^ and compared them to the CMV proteome using HEX, generating a list of melanoma-associated peptides highly similar to CMV (Supplementary Table S2). In the one patient, of whom we had available PBMCs, cross-reactivity between tumor and CMV peptides was assessed by ELISpot. Interestingly we observed that in this responder patient, PBMCs were significantly stimulated by both tumor peptides and their CMV homologous counterparts, suggesting that the CMV infection could have expanded viral T cell clones that could attack and kill tumor cells making seropositive patients more prone to react towards melanoma specific epitopes similar to CMV (Figure 7 A, B).

**Figure 7.**
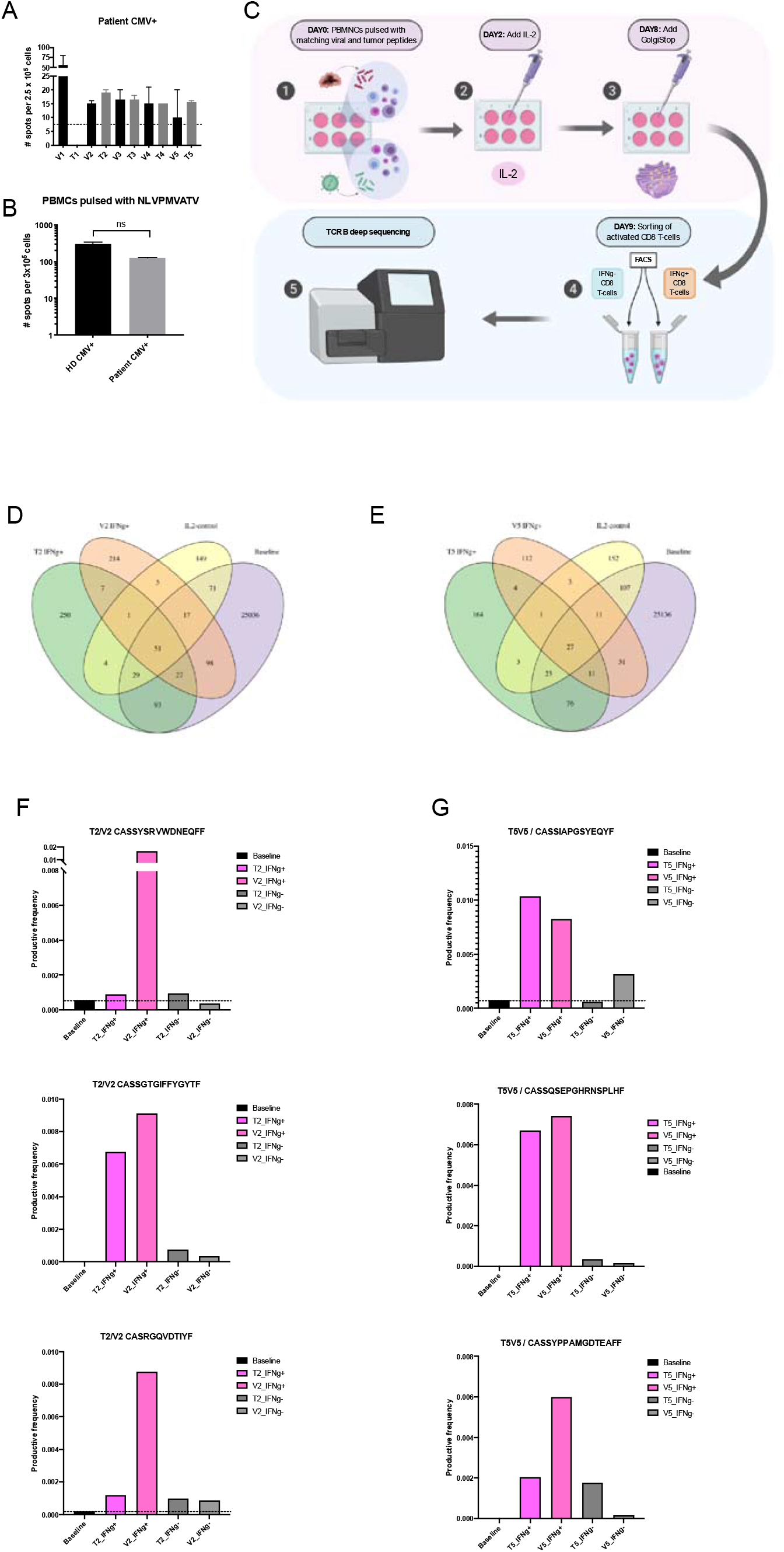
T-cell cross-reactivity between viral and tumor antigens shares high degree of homology. PBMCs derived from an HLA-A*02:01 patient with high serum level of anti-CMV Ab were pulsed with the peptides in Supplementary Table S2. The level of IFN-γ secreted by activated CD8+ T cells was detected by ELISpot assay. The dotted line indicates the noise level coming from unspecific activation of CTL in the negative control CMV resembling tumor (T1–5) and viral (V1–5) peptides presented in Supplementary Table S2 **(A)**. PBMCs derived from an HLA-A*02:01 positive patient with high serum level of anti-CMV Ab and from a healthy donor (HD) found positive for CMV response were tested for anti-CMV response by ELISpot assay using CMV-specific HLA-A*02:01 restricted peptide NLVPMVATV. P value was calculated using t-test with Mann-Whitney correction **(B)**. Experimental design for cross-reactivity assay. On day 0 primary human PBMCs were in vitro stimulated with matching viral and tumor peptides, on day 2 IL2 was added to the culture in order to sustain the primary cell culture until day 8 when GolgiStop was added. After inhibition by GolgiStop on day 9 the IFN-γ positive and negative fractions of CD8 T cells were sorted using FACS for further TCRβ sequencing **(C)**. Venn diagrams indicating the number of shared clones between viral or tumor antigen expanded active CD8+ T-cells and the baseline CD8+ T-cells. Clones that expanded with IL2 stimulation only were excluded from further analysis **(D&E)**. The top three clones with highest combined productive frequency among IFN-γ producing CD8+ T cell pool that overlapped between viral and tumor antigen expansion, but were not expanded with IL2 stimulation only. Bars represent the productive frequency of clones in IFN-γ producing (pink) and IFN-γ negative (grey) CD8+ T-cells after viral (striped fill) and tumor (solid fill) antigen expansion. Black dotted line represents the baseline expression (black bar) **(F&G)**.

This example indicates that molecular mimicry could explain part of the response of this patient but does not yet demonstrate involvement of cross-reactive T cells. In fact, a CMV positive cancer patient could theoretically expand two different T cell clones, one against the tumor peptide and one against the viral CMV peptide. To overcome this problem and finally assess whether homologous peptides could also activate and expand the same T cells clones we designed an additional cross reactivity experiment using healthy donors seropositive for CMV. In these individuals we investigated whether the same T cell clones were expanded when PBMCs were pulsed with viral (V) and tumor (T) peptides with high molecular mimicry. Thus, we activated PBMCs collected from a CMV seropositive donor, using these previously evaluated viral and tumor antigen pairs (named V2/T2 and V5/T5). After the activation, we sorted the IFN-γ positive and negative CD8^+^ T cells that were further analyzed with TCRβ sequencing (workflow of the experiment is presented in Figure 7C). From the TCRβ data we identified clones that were present in both activated T cell pools, pulsed either with viral or tumor peptides, but not present when stimulated with IL2 only (Figure 7 D&E). Following, among these shared clones, we filtered the top three clones based on the sum of productive frequency among the activated T cell pools and compared the productive frequency of these clones in each experimental T cell pool. Interestingly we observed that peptides with high molecular mimicry (V2/T2 and V5/T5) expanded the same T cell clones when compared to the baseline productive frequency and the productive frequency in IFN-γ negative CD8+ T-cell pool (Figure 7 F,G), demonstrating that homologous peptides predicted by HEX could be recognized by the same TCR. These findings indicate, for the first time, that molecular mimicry plays a role in mediating anti-tumor immune response and can potentially be exploited to design more efficient anti-cancer immunotherapies based on the pre-existing immunity of the patient.

## Discussion

In this study we present a novel bioinformatic tool (HEX) to identify tumor-specific MHC-I restricted peptides with high similarity to viral-specific peptides. Using peptides identified by this tool, we show that, in murine tumor models, viral pre-existing immunity enhances the efficacy of cancer immunotherapy via molecular mimicry. We further report, that in a cohort of human melanoma patients, with high humoral response to CMV, molecular mimicry between CMV- and tumor-antigens potentially plays a role in the response to checkpoint inhibitor therapy (anti-PD1), via activation of cross-reactive T-cells. In support of this, we also demonstrate that, in healthy donors with pre-existing immunity to CMV, melanoma-specific peptides similar to CMV-specific peptides activate and expand the same T cell clones. Our findings demonstrate that viral molecular mimicry favorably modifies the tumor immune-microenvironment, improving efficacy of immunotherapy.

The TCR is a highly promiscuous receptor and thus allows T-cells to recognize a very large variety of different targets^1, 2^. The idea that an immune response toward certain pathogens could lead to cross recognition of self-antigens is well established in the field of auto-immune diseases, a mechanism known as “molecular mimicry”^3^. Here, we hypothesized that a similar mechanism could also drive the anti-tumor immune response via cross-reactive T-cells.

Some studies have speculated that pathogen-specific cross-reactive T-cells could be responsible for extraordinary anti-tumor immune responses and prolonged PFS observed in some cancer patients^4, 8^. Although intriguing, this hypothesis has not been systematically studied nor has it impacted clinical practice, partly due to the lack of a proper tool to identify molecular mimicry between tumor and pathogen antigens. To overcome this problem, we developed a unique and novel software to rapidly identify tumor-specific MHC-I restricted peptides with high homology to pathogen derived MHC-I restricted peptides (HEX; Homology Evaluation of Xenopeptides). Contrarily to a simple alignment of sequences, this software assesses physical-chemical similarity at molecular level between an input sequence (single peptide or a list of peptides) and a list of pathogen-derived antigens^16^ and provides a positionally weighted^17^ alignment score to prioritize similarities occurring in the central section of the peptide, primarily involved in the interaction with the TCR^18^. Moreover, to increase the chances of identifying naturally presented epitopes, it predicts the MHC-binding affinity of both the input and the cognate viral peptides using state of the art MHC-binding-affinity predictors (NetMHC-API from IEDB servers)^19^.

We found that preimmunization with viral peptides similar to well characterized tumor peptides^10, 11, 12, 13^, efficiently slowed down the growth of subcutaneously injected melanoma tumor cells in mice, indicating that the pre-exposure to viral-derived peptides can affect the tumor growth. Rosato et al. have reported that virus-specific memory T cells can populate the tumor, enhancing checkpoint blockade therapy^9^. Herein, we propose the underlying mechanism to be molecular mimicry between viral and tumor antigens. We also observed that the tumor-homologous viral-derived peptides have therapeutic effect on already established tumors, indicating that a viral infection occurring during tumor progression could influence tumor response to treatment. Interestingly, mice intratumorally injected with the peptide-uncoated virus didn’t show any anti-tumor effect in contrast to what has been shown by Newman et al.^20^. On the contrary, viruses coated with highly homologous peptides produced a marked anti-tumor response showing that the effect was antigen-specific and based on molecular mimicry. The apparent discrepancy between these findings could be explained by the use of different viruses, with differing antigen repertoire and different degree of homology between virus and tumor, in the two studies. Tumor regression following viral administration or natural infection has previously been reported. This could be due to a generic mechanism, such as the “adjuvancy” effect of the virus with consequent recruitment of T cells at the tumor site, or to a more specific mechanism of shared antigens and activation of cross-reactive T-cells. Many coincidences have to occur to trigger potent mimicry-mediated tumor regression spontaneously, and, as for autoimmune diseases, the mimicry-mediated mechanisms may not be enough to trigger a biologically relevant T cell immune response, without other concomitant circumstances^3^. However, we envisioned that with the right tool to identify optimal homologous antigens, the mimicry between tumor and pre-existing exposure to a certain pathogen could be exploited to specifically boost the efficacy of active immunotherapy. Our findings potentially have important implications in peptide-selection for cancer vaccines, where peptides able to recruit antiviral-memory T cells in the tumor microenvironment might improve the outcome of treatment^4, 8^.

Recently, several attempts have been made to repurpose existing anti-viral T cells or using general vaccines to boost and/or redirect T cells against the tumors; Rosato et al.^9^ have used viral derived peptides to engage pre-existing T cells, Newman et al.^20^ have used direct intra-tumoral administration of the flu vaccine and Tähtinen & Feola et al. have previously used tetanus-derived peptides to engage tetanus-specific CD4+ T cells in pre-vaccinated tumor-bearing mice^21^. All these approaches have been successful in preclinical models and some of them have also proceeded to clinical testing. We suggest that they could be even more efficient if designed based on molecular mimicry with regards to the patients’ pre-existing immunity.

We also find that molecular mimicry could partially explain the efficacy of ICPIs in human cancer patients. Novel ICPIs, such as anti-PD1, have significantly improved the survival of patients with solid tumors, especially in metastatic melanoma, when compared to other commonly used therapies such as radiation and chemotherapies^22, 23^. However, despite the enhanced survival and efficient response rates, it is yet unknown why some patients benefit more than others. Certainly, the direct administration of viruses to the tumor site enhances the influx of T cells predisposing the ICPIs to work more efficiently^24, 25^. However, the link between these viral T cells and anti-tumor response is still unknown. Molecular mimicry could be the missing link, at least in some cases, explaining why anti-viral T cells at the tumor site have such a beneficial effect. Therefore, we studied a cohort of metastatic melanoma patients undergoing anti-PD1 therapy and measured their serum anti-CMV and anti-EBV IgG titers, as these are common viruses and specific T cells have been often found in the tumors of patients^26^. In particular CMV reactivations have previously been reported in ICPI treated patients^26^ and its association to beneficial therapy response has been speculated. Our results indicated that the patients with high titer of CMV-specific IgG levels had significantly longer PFS. On the contrary, the EBV-specific IgG levels did not associate with prolonged PFS. Interestingly, no significant differences could be observed in the general immunization between the responder and non-responder cohorts supporting the hypothesis that the immune response specific to CMV infection, might contribute to the prolonged survival in anti-PD1 treated melanoma patients. To further strengthen these findings, we observed that the PBMCs, from a patient with high anti-CMV-IgG titer, reacted with melanoma antigens similar to CMV peptides. In addition to this, a deeper evaluation on the cross-reactivity by deep TCRβ sequencing indicated that, indeed, the same TCR clones were expanded in the activated T-cell populations when pulsed with mimicking viral- and tumor-derived antigens even when absent at the baseline. This data indicates the possibility of molecular mimicry, between viral- and tumor-derived antigens, to contribute to cancer-specific T-cell immunity. Thus, based on our *in vitro* cross-reactivity results, together with the association of high serum CMV IgG levels and prolonged survival of melanoma patients, we also believe that the molecular mimicry between CMV and melanoma could provide a clinical advantage for these patients undergoing ICPI therapy.

We are aware of some limitations of our study. Our HEX tool only predicts binding affinity of CD8 restricted epitopes. These were, however, prioritized as CD8 T cells are the ones mainly involved in recognizing both virally infected and transformed cells. Secondly, HEX does not consider whether the peptides are naturally processed and this could affect the number of false positive candidate peptides^27^. The small patient cohort available for this study is an additional limiting factor. Thus, in order to explore molecular mimicry in a clinical setting, further studies with larger patient cohorts in controlled clinical trials together with other possible target epitopes is warranted.

The results of this study indicate that viral infections could have an impact on tumor growth and clearance. Additionally, we show cross-reactivity of cytotoxic T-cells against viral-and homologous tumor-derived antigens, selected using our novel software HEX. Our findings highlight the importance of viral pre-existing immunity in cancer immunotherapy, and suggest the use of HEX to select highly homologous viral-tumor antigens to engage cross-reactive T cells.

## Materials and Methods

### HEX (Homology Evaluation of Xenopeptides)

Homology Evaluation of Xenopeptides (HEX) is a novel in silico platform that compares similarity between tumor peptides (reference peptides) and viral peptides (query peptides). It utilizes several metrics in order to expediate candidate peptide selection. This is done by incorporating both novel methods (peptide scoring and alignment scoring algorithm) and integrated pre-existing methods (MHC-I binding prediction). HEX comes with a number of precompiled databases of known proteins, such as proteins derived from viral pathogens and the human proteome^16^.

Peptides are ranked by a score (henceforth, B-score) as previously described^16^, which represents the log-likelihood of the viral peptide being recognized by a T-cell. The associated scoring matrix is generated ad hoc based on the amino acid composition of the reference peptide, as opposed to experimentally. In particular, in the matrix rows representing the amino acid position in the peptide and columns representing each of the 20 standard amino acids, amino acid positions of the reference peptide were assigned the same high score and other positions were assigned the same low score. The B-score is the agonist log-likelihood score for this special matrix and is given by:

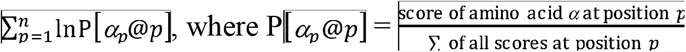

Alignments are computed pairwise between peptides in the query set against the reference set. For a given pair of peptides, their alignment is calculated by the summing the distance scores between pairs of amino acids in the same position. Scoring is weighted to prioritize similarity between more central amino acids in the peptide. HEX supports both BLOSUM and PAM substitution matrices across several evolutionary distances.

MHC class I binding affinity predictions are made using NetMHC^19^ via the Application programming interface of IEDB (http://tools.iedb.org/main/tools-api/) and are then parsed and collated within the tool. The user can specify their desired scoring method or return a number of recommended results. Predictions for a number of human and murine MHC-I alleles are supported.

Users are able to select peptides by their own criteria or allow peptides to be selected by a random forest model. The random forest was trained on experimental outcomes of peptides chosen by the authors. Feature importance was determined by out-of-bag (OOB) increase in mean squared error (MSE) and cross-validated on an unseen sample of the peptides. HEX was developed as a web application using the R package Shiny and is accessible at https://picpl.arcca.cf.ac.uk/hex/app/ without user registration. The source code is available at https://github.com/whalleyt/hex.

### Patients and samples

In total of 16 stage four metastatic melanoma patients were treated with anti-PD1 monoclonal antibody in the Helsinki University Central Hospital (HUCH) Comprehensive Cancer Center. Patients were randomly selected to receive either nivolumab (n = 7) infusions every second week or pembrolizumab (n = 9) infusions every third week. The study was approved by the HUCH ethical committee (Dnro 115/13/03/02/15). Written informed consent was received from all patients and the study was conducted in accordance with the Declaration of Helsinki. For detailed patient characteristics see Supplementary Table S3.

Peripheral blood samples (3ml EDTA blood, 50ml Heparin blood) were collected from three time-points; before initiation of treatment, after one and three months of treatment. From these the plasma was separated by centrifuging and then stored in –70C°. The CMV and EBV IgG levels were measured from thawed EDTA plasma samples using VIDAS CMV IgG (BioMérieux, Marcy-l’Etoile, France) and Siemens Enzygnost Anti-EBV/IgG kits (Siemens Healthcare Diagnostics, Marburg, Germany). The immunoglobulins (IgA, IgM, IgG) from thawed Heparin plasma were measured in the central laboratory of the Helsinki University Central Hospital (HUSLAB).

### Cell lines and human samples

The murine melanoma cell line B16-F10 was purchased from the American Type Culture Collection (ATCC; Manassas, VA, USA). Cells were cultured in RPMI (Gibco, Thermo Fisher Scientific, US) with 10% fetal bovine serum (FBS) (Life Technologies), 1% Glutamax (Gibco, Thermo Fisher Scientific, US), and 1% Penicillin and Streptomycin (Gibco, Thermo Fisher Scientific, US) at 37°C/ 5% CO_2_.

The cell line B16-OVA, a mouse melanoma cell line modified to constitutively express chicken Ovalbumin (OVA), was kindly provided by Prof. Richard Vile (Mayo Clinic, Rochester, MN, USA). These cells were cultured in RPMI Low glucose (Gibco, Thermo Fisher Scientific, US) with 10% FBS (Gibco, Thermo Fisher Scientific, US), 1% Glutamax, 1% Penicillin and Streptomycin (Gibco, Thermo Fisher Scientific, US) and 1% Geneticin (Gibco, Thermo Fisher Scientific, US) at 37°C/ 5% CO_2_.

All cells were tested for mycoplasma contamination with a commercial detection kit (Lonza – Basel, Switzerland). Isolated human PBMCs were frozen in FBS supplemented with 10% DMSO and then maintained in liquid nitrogen until use.

Cryopreserved PBMCs were thawed and rested overnight at 37°C/ 5% CO_2_ in complete RPMI medium supplemented with 10% FBS, 1% Glutamax, 1% penicillin-streptomycin over night before plating them for ELISPOT.

### Peptides

All the peptides used in this study were purchased from Zhejiang Ontores Biotechnologies Co. (Zhejiang, China) or GenScript (New Jersey, USA) 5mg > 90% purity. The sequences of all the peptides used in this study are found in Supplementary Table S1 and S2.

### PeptiCRAd preparation

All PeptiCRAd complexes described in this work were prepared by mixing Adenoviruses and polyK-tailed peptides according to the following protocol: 1×10^9^vp were mixed with 20ug peptides; after vortexing, the mixture was incubated at room temperature for 15 min; successively, PBS was added up to the injection volume (50uL). For the TRP2-PeptiCRAd, 1×10^9^vp were mixed with 20ug of 6K-TRP2_180–188_ peptide, while the Viral-PeptiCRAd was prepared using 1×10^9^vp mixed with 5ug of each viral 6K-peptide homologous for TRP2_180–188_. gp100 PeptiCRAd was prepared using 1×10^9^vp mixed with 20ug of 6K-GP100_25–33_ peptide.

New PeptiCRAds were prepared before each experiment using fresh reagents. All dilutions of virus and peptides required before incubation were performed in sterile PBS. Viruses were generated, propagated, and characterized as elsewhere described^14^.

### Animal experiments and ethical permits

All animal experiments were reviewed and approved by the Experimental Animal Committee of the University of Helsinki and the Provincial Government of Southern Finland. All in vivo models were carried using C57BL/6JOlaHsd mice obtained from Scanbur (Karlslunde, Denmark).

For the first animal experiment, 8- to 9-week-old immune competent female C57BL/6J mice were divided in 4 groups. N = 3 mice were used as mock group, n = 7 mice were used to form each of the three different treatment groups. Each group was immunized with a different pool of viral-derived peptides homologous to tumor epitopes. Mice were vaccinated twice and injections were performed at one-week interval (day 0 & 7) at the base of the tail with 40ug of peptides pool (10ug each peptide) and 40ug of adjuvant (VacciGrade poly(I:C) – Invivogen) in a final injectable volume of 100ul. Naïve mice (PBS injected) were used as Mock group. At day fourteen mice were injected with 3*10^5^ B16-OVA cells on the right flank and tumor growth was followed until endpoint was reached.

For the treatment of established tumors, we tested 2 different tumor cell lines: B16-OVA cells and the more aggressive B16F10 cells. 3*10^5^ B16-OVA cells and 1*10^5^ B16-F10 were injected subcutaneously on the right flank of 8- to 9-week-old immune competent female C57BL/6J mice. Successively, these mice were randomly divided in 4 groups of 7–8 mice for each tumor cell line. A mock group was treated with PBS; a second group was treated with uncoated Adenovirus; a third group was treated with Adenovirus coated with TRP2_180–188_ (TRP2–PeptiCRAd); the last group was treated with Adenovirus coated with TRP2-homologous viral peptides (Viral–PeptiCRAd).

Mice were intratumorally treated twice and injections were performed at two days interval (day 10 and 12 from tumor engraftment) a final volume of 50uL per injection. Tumor growth was measured every 2 days with a digital caliper until endpoint was reached. The tumor volume was mathematically calculated according to the following formula:

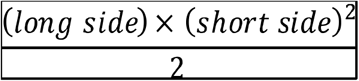

The median of the tumor volume measurement of the last day identified the therapeutic success threshold and showed as a dotted line in the graphs. Mice that at the end point showed a tumor volume below the threshold were considered responders, while mice above it were considered non-responders.

As for the animal experiment depicted in Figure 5, we used a combination of all the above-mentioned methods. Half of the cohort of C57BL/6J female mice (8- to 9-week-old) was preimmunized with the pool of viral-derived peptides similar to TRP2 (Supplementary Table S1) according to the protocol described previously. After two weeks from the first immunizations all the mice were subcutaneously injected with 3*10^5^ B16-OVA cells on their right flank. When the tumor was visible, mice were divided into 6 groups of n = 8–10 mice per group, and were intratumorally injected three times (two days apart) with either PBS (Mock group), gp100-coated PeptiCrad (gp100 group) or TRP2-coated PeptiCrad (TRP2 group). Tumor growth was followed as previously described until the endpoint of the experiment was reached.

### ELISpot assay

To assess the amount of active antigen specific T-cells, interferon-γ (IFN-γ) secretion was measured by ELISPOT assay from IMMUNOSPOT (CTL, Ohio USA) for the murine IFN-γ and MABTECH (Mabtech AB, Nacka Strand, Sweden) for the human IFN-γ

Fresh mice splenocytes collected at the end point of the experiment have been used. The procedure was carried out according to the manufacturer instructions. In brief, for murine IFN-γ, 3*10^5 of splenocytes/well were plated at day 0. Cells were stimulated with 2ug/well of peptides. After 3 days of incubation at 37°C/ 5% CO_2_, plates were developed according to the kit’s protocol.

For human IFN-γ ELISPOT, human PBMCs were thawed and let rest over-night at 37°C/ 5% CO_2_ in complete medium. The following day, 3×10^5^ PBMCs/well were plated and stimulated with 2ug/well of peptides. After 48h of incubation at 37°C/ 5% CO_2_ the plates were developed according to the manufacturer’s protocol. Plates were sent to CTL-Europe GmbH to be analyzed.

### Flow cytometry analysis

Intracellular staining was performed using FOXP3 Fixation/Permeabilization Buffer (Biolegend - 421403) according to the manufacturer instructions.

Antibodies used in this study were the following: TruStain fcX (anti-mouse CD16/32) (Biolegend - 101320), CD3-BV711 (BD - 563123), CD4-PECF594 (BD - 562285),

CXCR3-APC (BD - 562266), CD44-V450 (BD - 560451), Ki67-PECy7 (BD - 561283), CD8a (KT15)-FITC (Proimmune 1705F/33790).

The data were acquired using BD LSR-FORTESSA flow cytometer and subsequently analyzed using FlowJo software v10.

### Cross-reactivity assay

The cross-reactivity of T-cells against the viral and tumor antigens sharing high similarity was studied by first expanding the PBMCs from CMV seropositive donor in vitro using the matching viral and tumor antigens (V2/T2 and V5/T5).

Cryopreserved PBMCs were thawed in warm 37°C RMPI and then rested overnight at 37°C, 5% CO_2_ in complete RPMI medium supplemented with 10% FBS, 1% L-glutamine, 1% penicillin-streptomycin, before stimulation with viral or tumor antigens. After resting overnight, on day 0 the PBMCs were plated as 6×10^6^ cells/well on 6-well plates with the selected peptides in final concentration of 4 µM/ml and incubated 48 hours at 37°C, 5% CO_2_ in complete RPMI medium, duplicate samples were used for each peptide stimulation. NLV peptide was used as a positive control, for negative control no peptide stimulation was used. On day 2 half of the media was replaced with fresh complete RMPI with IL2 in final concentration of 20 IU/ml. On day 8 GolgiStop^TM^ (BD BioSciences) was added according to manufacturer’s instructions and cells were incubated overnight before cell sorting.

On day 9 the cells were collected and stained with surface markers; CD3, CD8, CD45, CD56 and NLV pentamer followed by fixing and permeabilization using BD Cytofix/Cytoperm™ (BD BioSciences) and staining with intracellular IFN-γ. The IFN-γ positive and negative CD8^+^ T-fractions were sorted for DNA extraction followed by TCR β deep-sequencing (FIMM). The workflow of cross-reactivity assay is illustrated in Figure 7C.

## Data Availability

All the data presented in this manuscript are available

## Supplementary Materials

**Supplementary Table S1:**
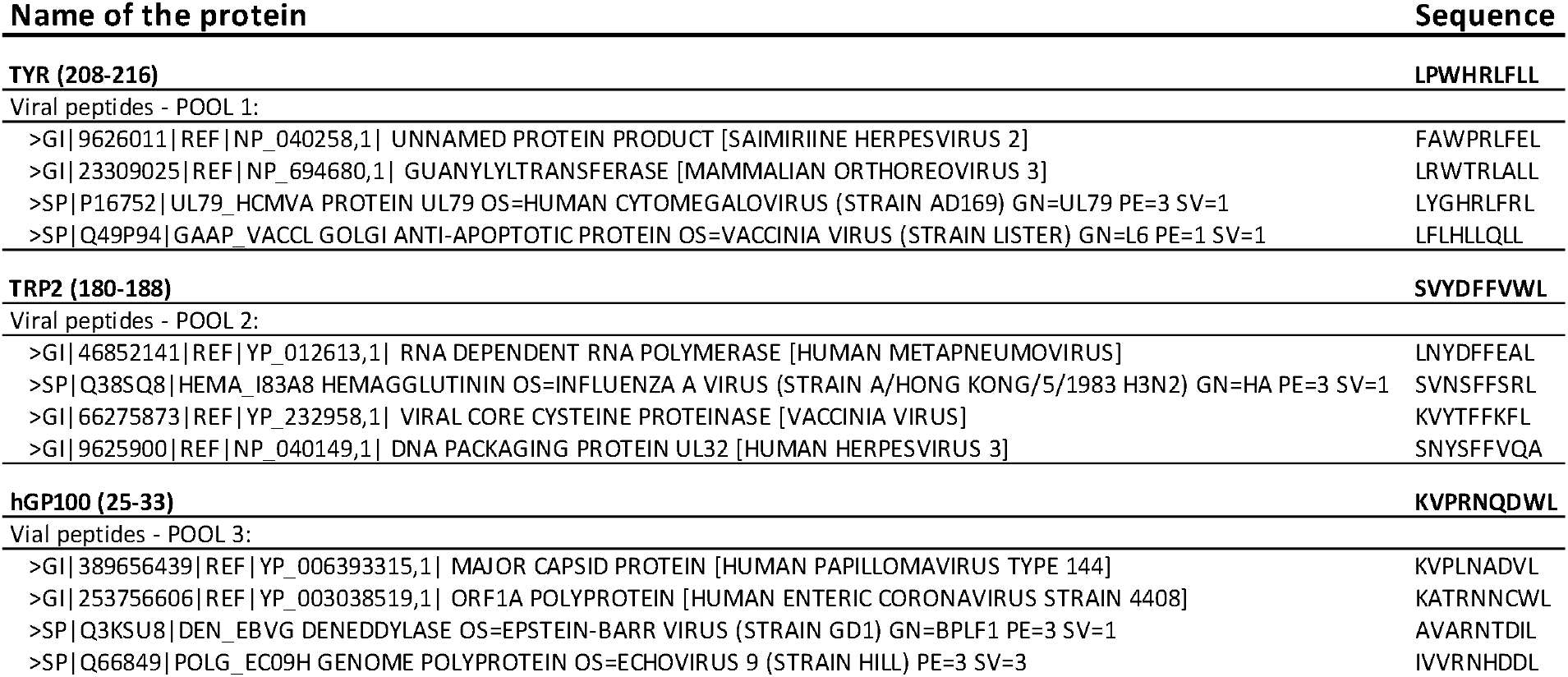
Peptides used for the animal experiment. Known melanoma tumor peptides (TRP2_180–188_, hGP100_25–33_ and TYR_208–216_) were analyzed with HEX. The best viral candidate peptides proposed by the software were selected and a pool composed by the best 4 viral-derived peptides per each original tumor epitope was tested *in vivo*.

**Supplementary Table S2:**
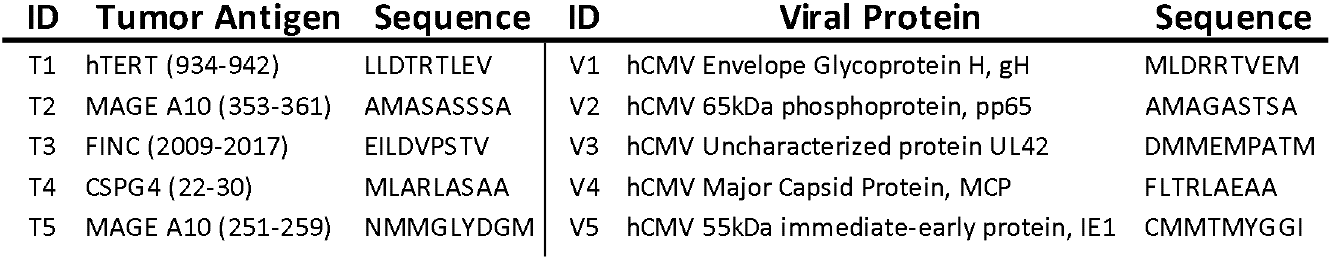
Peptides used for the ELISpot on patients’ PBMCs. Known Melanoma-associated antigens were analyzed with HEX. The best human CMV-derived candidate peptide per each antigen was chosen to be tested in vitro.

**Supplementary Table S3:** Patient characteristics. The Eastern Cooperative Oncology Group (ECOG) performance status1. Stage classification, AJCC7 Metastasis stage2:M1a = metastasis in distant skin sites or areas under the skin or in distant lymph nodes with normal LDH, M1b = metastasis in lung with normal LDH levels, M1c = metastasis in internal organ or any other metastasis with elevated LDH. Treatment responses evaluated using the RECIST-criteria3; Best achieved response during anti-PD1 therapy: CR = complete response, PR = partial response, SD = stable disease, PD = progressive disease. Response status; Responder (CR, PR or SD > 6mo), non-responder (SD < 6mo, PD). Pretreatment anti-CMV-IgG; High ≥ 93 UA/ml < Low.

| No | Age at initiation | Sex | ECOG | Stage | Metastasis stage | Brain metastasis | Drug | BestResponse | FollowUp (months) | PFS (months) | Response status | Pretreatment anti-CMV-IgG |
| --- | --- | --- | --- | --- | --- | --- | --- | --- | --- | --- | --- | --- |
| 1 | 71 | 1 | 0 | 4 | M1b | no | Pembrolizumab | SD | 27 | 9 | Responder | High |
| 2 | 78 | 1 | 0 | 4 | M1c | no | Pembrolizumab | PR | 21 | 20 | Responder | High |
| 3 | 70 | 1 | 1 | 4 | M1c | no | Nivolumab | PD | 20 | 4 | Non-responder | High |
| 4 | 84 | 2 | 0 | 4 | M1a | no | Nivolumab | CR | 18 | 17 | Responder | High |
| 5 | 78 | 1 | 0 | 4 | M1c | no | Pembrolizumab | PD | 12 | 4 | Non-responder | High |
| 6 | 66 | 2 | 0 | 4 | M1c | no | Nivolumab | PR | 12 | 11 | Responder | High |
| 7 | 59 | 2 | 1 | 4 | M1c | no | Nivolumab | PD | 27 | 6 | Non-responder | Low |
| 8 | 53 | 1 | 0 | 4 | M1c | no | Nivolumab | PD | 26 | 1 | Non-responder | Low |
| 9 | 75 | 1 | 0 | 4 | M1b | no | Pembrolizumab | SD | 22 | 6 | Responder | Low |
| 10 | 63 | 1 | 1 | 4 | M1b | no | Pembrolizumab | PD | 21 | 2 | Non-responder | Low |
| 11 | 69 | 1 | 0 | 4 | M1a | no | Pembrolizumab | SD | 21 | 8 | Responder | Low |
| 12 | 66 | 2 | 0 | 4 | M1b | no | Pembrolizumab | PR | 20 | 3 | Responder | Low |
| 13 | 69 | 2 | 0 | 4 | M1c | no | Pembrolizumab | PR | 21 | 18 | Responder | CMV neg |
| 14 | 70 | 1 | 1 | 4 | M1c | no | Nivolumab | PR | 17 | 16 | Responder | CMV neg |
| 15 | 79 | 2 | 0 | 4 | M1a | no | Nivolumab | PR | 18 | 12 | Responder | CMV neg |
| 16 | 74 | 1 | 0 | 4 | M1a | no | Pembrolizumab | CR | 22 | 20 | Responder | no data |

1. Oken M, Creech R, Tormey D, et al. Toxicity and response criteria of the Eastern Cooperative Oncology Group. *Am J Clin Oncol.* 1982;5:649-655 2. Edge SB, Compton CC. The American Joint Committee on Cancer: the 7th edition of the AJCC cancer staging manual and the future of TNM. *Ann Surg Oncol.* 2010;17:1471-4. 3. Eisenhauer EA, Therasse P, Bogaerts J, Schwartz LH, Sargent D, Ford R, et al. New response evaluation criteria in solid tumours: revised RECIST guideline (version 1.1). *Eur J Cancer.* 2009;45:228-47.

**Supplementary Figure 1:**
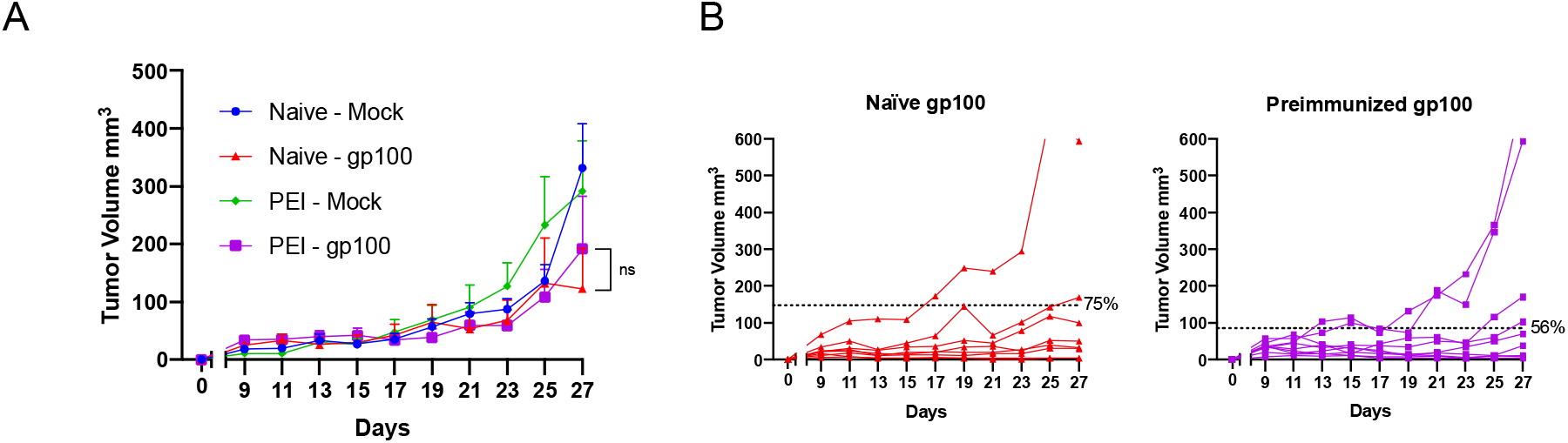
Average tumor growth shown as mean ± SEM. Statistical analysis Two-way ANOVA (**A**). Tumor volume curves of individual mice per each group. The dotted line identifies the threshold of the therapeutic success rate and represents the median of the normalized volumes measured at the endpoint for the Naïve groups and the PEI groups respectively (**B**)

## Acknowledgments

We thank all the co-authors for their support and hard work and to the Advanced Research Computing at Cardiff (ARCCA) for hosting and assisting with HEX. The authors would like to thank also the patients, clinicians and study-nurses for their participation.

## Funding

This work has been supported by European Research Council under the European Union’s Horizon 2020 Framework programme (H2020)/ ERC-CoG-2015 Grant Agreement n.681219. Moreover, this research was supported by Helsinki Institute of Life Science (HiLIFE), Jane and Aatos Erkko foundation and Cancer society of Finland. Additionally, this work has been supported by Finnish Cancer Organizations, Sigrid Juselius Foundation, Relander Foundation, State funding for university-level health research in Finland, Fican South funding and HiLife fellow funds from the University of Helsinki.

## Author contributions

J.C.: acquisition, analysis and interpretation of data, drafting of the manuscript; H.H.E.K.: acquisition, analysis and interpretation of data, writing of the manuscript; T.W.: coding of the software, development and maintenance; C.C.: conception and design of the study, acquisition, analysis and interpretation of data, drafting of the manuscript; M.G.: interpretation of data and revision of the manuscript; S.F.: interpretation of data and revision of the manuscript; K.D.P.: interpretation of data and revision of the manuscript; F.S.H.: interpretation of data and revision of the manuscript; M.M.H.: treating clinician, provided the clinical data and samples from patients; S.M.: treating clinician, provided the clinical data and samples from patients; B.M.: acquisition, analysis and interpretation of data; M.F.: acquisition, analysis and interpretation of data; A.S.K.: conception and design of the study, analysis and interpretation of data, drafting of the manuscript; S.M.M.: conception and design of the study and interpretation of data, drafting of the manuscript; B.S.: conception and design of the study, analysis and interpretation of data, drafting of the manuscript; V.C.: conception and design of the study, analysis and interpretation of data, drafting of the manuscript.

## Competing Interests

J.C., H.H.E.K., T.W., C.C., M.G., S.F., K.D.P., F.S.H., M.M.H., S.M., B.M., M.F., A.S.K. and B.S. declare to not have any conflict of interests. S.M.M. has received honoraria and research funding from Novartis, Pfizer and Bristol-Myers Squibb (not related to this study). V.C. is co-founder and shareholder of the Valo Therapeutics LTD.

## Data and materials availability

All data associated with this study are available on request from the corresponding author.

## Notes

### Author Declarations

The study was approved by the HUCH ethical committee (Dnro 115/13/03/02/15). Written informed consent was received from all patients and the study was conducted in accordance with the Declaration of Helsinki.

